# COVID-19 epidemic scenarios into 2021 based on observed key superspreading events

**DOI:** 10.1101/2021.04.14.21255436

**Authors:** Mario Santana-Cibrian, M. Adrian Acuña-Zegarra, Carlos E. Rodríguez, Ramsés H. Mena, Jorge X. Velasco-Hernandez

**Affiliations:** CONACYT-Instituto de Matematicas UNAM, Queretaro, 76230, Mexico; Departamento de Matemáticas, Universidad de Sonora, Sonora, 83000, Mexico; Department of Probability and Statistics, IIMAS UNAM, CDMX, 01000, Mexico; Instituto de Matemáticas UNAM, Queretaro, 76230, Mexico

**Keywords:** Superspreading, SARS-CoV-2 epidemic, Kermack-McKendrick models, Epidemic projections, Bayesian inference

## Abstract

Key high transmission dates for the year 2020 are used to create scenarios of the evolution of the COVID-19 pandemic in several states of Mexico for 2021. These scenarios are obtained through the estimation of a time-dependent contact rate, where the main assumption is that disease behavior is heavily determined by the mobility and social activity of the population during holidays and other important calendar dates. First, changes in the effective contact rate on predetermined dates of 2020 are estimated. Then, this information is used to propose different scenarios for the number of cases and deaths for 2021. The fundamental assumptions behind this methodology are that the effective contact rate incorporates the main superspreading transmission events during last year, that each region has an independent epidemic not explicitly interconnected with other regions and, finally, that there are no new highly transmissible SARS-CoV-2 variants active during the timeline of the forecasts. Also, several levels of vaccination are considered to analyze their impact on the projections of the epidemic curve. The objective is to generate a range of scenarios that could be useful to evaluate the possible evolution of the epidemic and its likely impact on incidence and mortality.

## 1 Introduction

As a consequence of the COVID-19 epidemic’s impact on the regional and global economy, one of the most relevant problems that every country faces is to decide how and when businesses, public centers, tourism, schools, and universities can safely reopen [21]. Therefore, it is of the highest importance to postulate plausible scenarios for the evolution of the SARS-CoV-2 pandemic to design effective reopening strategies for decision-makers. This knowledge is even more pressing in countries that lack of infrastructure (i.e., testing, contact tracing) to acquire a more precise or, perhaps we should say, a less uncertain idea of the behavior of the pandemic on days or, ideally, weeks into the future.

During 2020, much effort was centered on projecting the fate of the COVID-19 pandemic and evaluating the efficacy of the mitigation strategies adopted to contain it [7, 14]. The transmission dynamics of the pandemic has been modeled using many different methodologies, several of them centered on estimating the effective reproduction number *R*_*t*_ or using some version of the well-known Kermack-McKendrick model [8, 15, 28, 33].

Around the world, nonpharmaceutical interventios (NPIs) vary in strength. Examples include lockdowns, use of face masks, regulations on the number of people allowed in meetings, and so forth. They range from strict and mandatory enforcement by the governments to a voluntary personal decision. The rationale behind any particular version of a mitigation strategy followed in each country or region is generally based on local or national conditions that intermix public health, economic and political factors and perspectives [4]. One factor, however, that has shown to be decisive in the success or failure of a given containment or mitigation strategy and that has show remarkable difficulty to control or project is human behavior. Recently, some efforts in modeling this topic have been reported [3].

In Mexico, states present different shapes of epidemic curves. For example, Fig. 1 shows the epidemic curves of Mexico City, Queretaro state, and Tamaulipas state until May 2021. There are some features in these curves worth describing. One is the initial slow growth of the epidemic that seems almost linear in Mexico City and Queretaro state with a shorter growth period in Queretaro state; the second is the long plateau that develops in Mexico City and a much shorter one in Queretaro state, and the third an epidemic curve with two peaks in Tamaulipas.

**Fig. 1.**
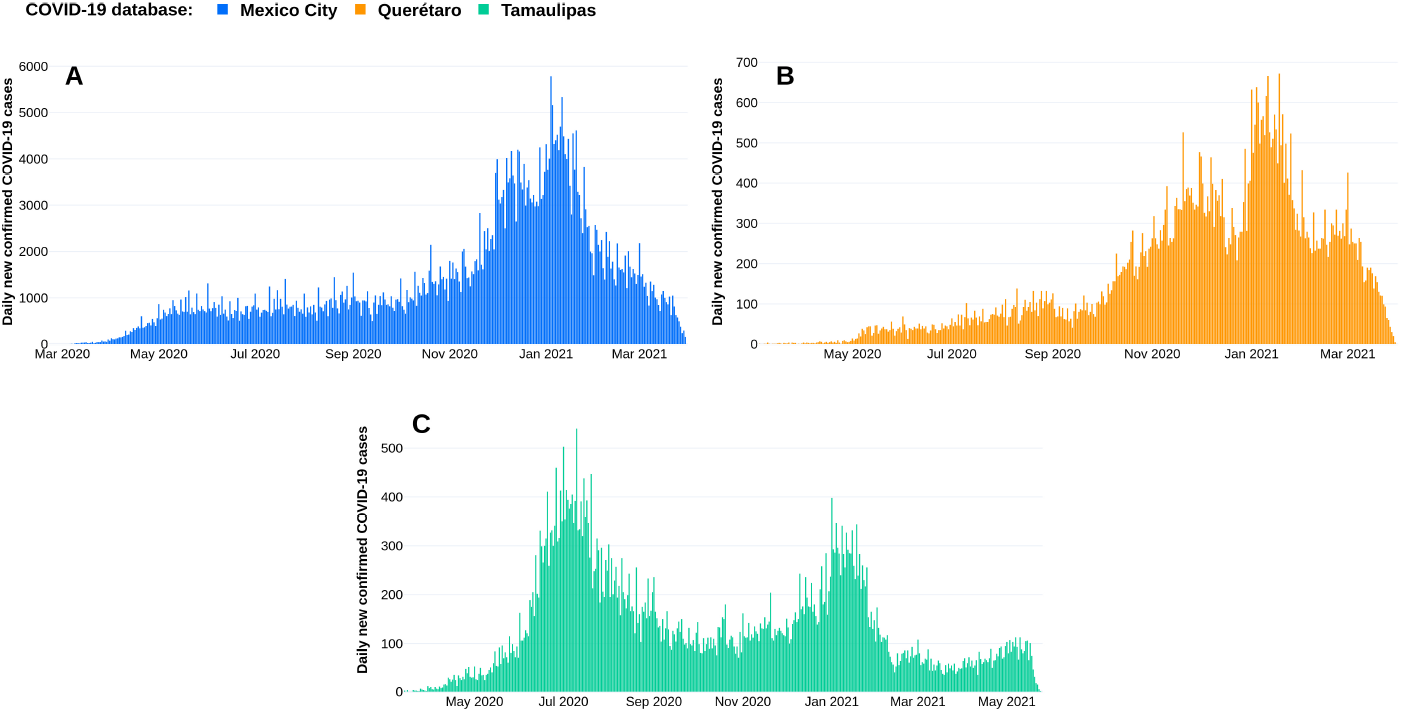
Daily number of COVID-19 confirmed cases by symptoms onset until May 24, 2021 for A) Mexico City, B) Queretaro state and C) Tamaulipas state.

The initial quasi-linear growth of the epidemic cannot be explained by classical models. Previously it has been have argued that the initial slow growth was a consequence of an early application of largely voluntary mitigation measures [1]. The first recorded case in Mexico occurred by the end of February 2020, and the first mitigation measures were applied 25 days later, on March 23, 2020. Moreover, there were superspreading events in Mexico City during Easter celebrations (April 6-12) and early May (April 30 to May 10) that shifted the day of maximum incidence to the end of May [1, 26], and pushed the epidemic into a quasi-stationary state characterized by values of *R*_*t*_ *≈* 1. This behavior, observed around the world, has been explored in [16, 27, 31]. In particular, [31] claim that this quasi-linear growth and the maintenance of the effective reproduction number around *R*_*t*_ *≈* 1 for sustained periods involves critical changes in the structure of the underlying contact network of individuals.

Superspreading events are indeed associated to heightened population mobility and social activity [2, 5, 9, 18, 24, 25, 29, 32, 34]. It is a phenomenon that has shown to be determinant for the time evolution of many infectious diseases [19]. The SARS-CoV-2 epidemic dynamic observed in Mexico City and, in general, in the country, has been driven by events associated with heightened mobility and increased social activities during holidays and other important calendar key dates. The characteristic plateau of Mexico City reached by the end of May, 2020, for example, has been explained by a succession of events that were essentially super-spreading events [1, 26].

Now that we have more than one year of data regarding the evolution of the COVID-19 pandemic, the following question arises: can we use the past history of the epidemic curve to project the main likely tendencies in the behavior of COVID-19 for the rest of 2021? The objective of the present work is to provide an answer to that question for the case of the Mexican epidemics. In order to do that, a characterization of the transmission dynamics of COVID-19 is presented based on the assumption that human behavior during specific dates is the main factor driving the evolution of the pandemic. It is postulated that a modification of the Kermack-McKendrick model with a time-dependent effective contact rate can accurately describe the observed number of confirmed cases and deaths of COVID-19. The effective contact rate will change in calendar dates associated with religious or civic holidays, commercial incentives and government interventions, when superspreading events or changes in tendency occurred. Then, this characterization of the transmission dynamics of the disease is used to develop a methodology to generate epidemic scenarios that use the known history of the disease recorded during 2020 and early 2021. The effect of these change points or key calendar dates on the contact rate occurs at a much faster scale than changes due to the implementation or lifting of NPIs [12, 17]. All these dates are known in advance, and activities can be planned (short vacations, family visits, among others). We focus on the epidemic developing in Mexico, taking as examples three particular states (Mexico City, Queretaro state and Tamaulipas state).

Our approach is similar to the one presented in [12]. However, to determine changes in the effective contact rate, besides looking at the actual effects of governmental mitigation measures, we look at particular events on dates related to civic, religious, and official vacation periods known in advance each year.

The paper is organized as follows. Section 2 presents the methodology used. Section 3 describes the results of the estimation process and the proyected scenarios. Finally, Section 4, contains the discussion about this work.

## 2 Methodology

This section describes the underlying mathematical model, the estimation of the time-varying effective contact rate and the basic assumptions for the projection of scenarios.

### 2.1 Mathematical model

A compartmental model is used to describe the evolution of the COVID-19 epidemic. Fig. 2 shows the corresponding model diagram. The model considers three classes of infected individuals: asymptomatic (*I*), symptomatic (*Y*), and reported (*T*). Once reported, infected individuals are effectively isolated and are no longer participants in the transmission process. The three infected classes can recover (*R*) but only reported symptomatic individuals can die (*D*) since we consider the ideal case where all severe cases are reported. Susceptible (*S*) individuals can be vaccinated (*V*) with an effective vaccination rate *ψ*. Both vaccinated and reported individuals will eventually return to the susceptible state after a certain, possibly different, immunity period. Vital dynamics are also included since this work is aimed to produce mid-term scenarios for the epidemic. The full set of equations is given by

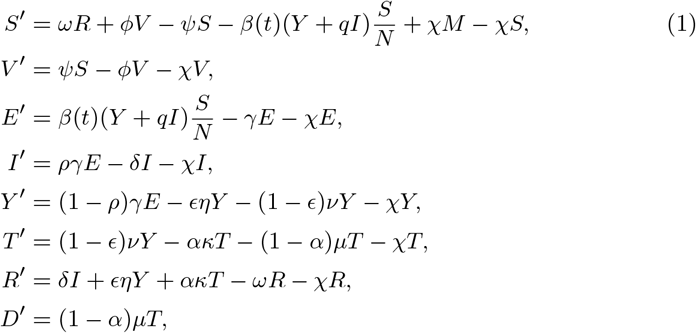

where *N* = *S* + *V* + *E* + *I* + *Y* + *R* is the population that participates in the infectious process and *M* = *N* + *T* is the total population at time *t*.

**Fig. 2.**
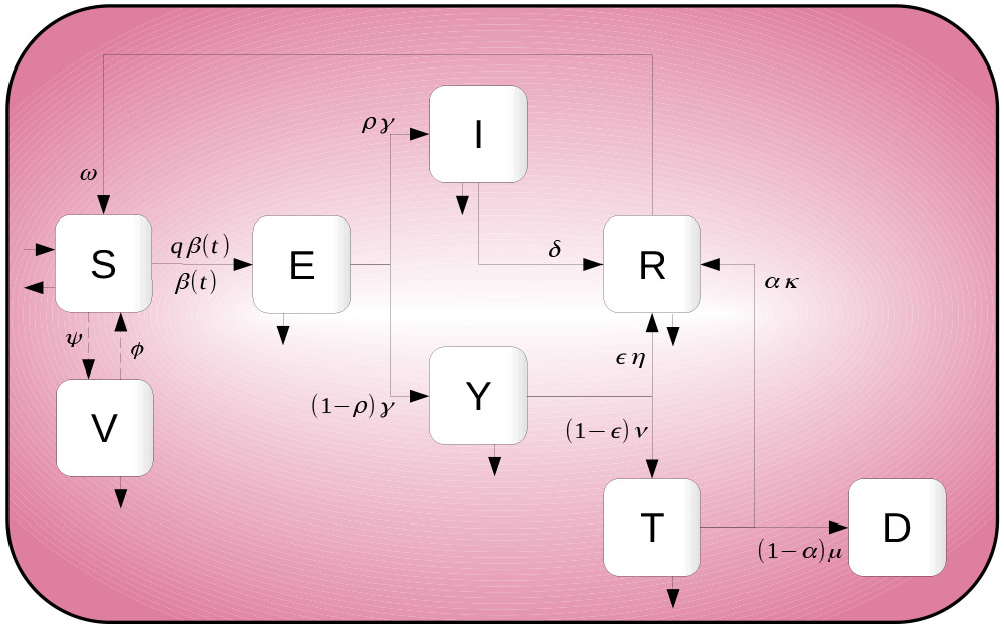
Diagram for the mathematical model. There are three types of infectious individuals: asymptomatic (I), symptomatic (Y) and reported (T), but this last group does not play a role in the transmission. The other state variables represent: susceptible (S), vaccinated (V), exposed (E), recovered (R) and dead (D).

The effective vaccination rate is deduced as

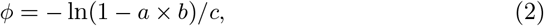

=
where *a* is the target coverage (% of the population targeted to receive the vaccine), *b* the vaccine efficacy and *c* the time horizon in which *a* should be achieved. In what follows *φ* defines plausible scenarios because in Mexico vaccination roll-outs are regionally heterogeneous and vaccine availability is limited.

The basic reproduction number is given by

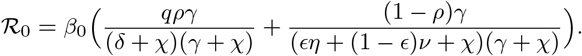

The parameter *β*_0_ is the effective contact rate at the start of the epidemic. Furthermore, the effective reproduction number is calculated as

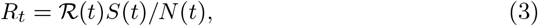

where

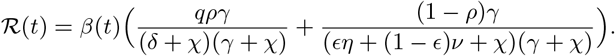

Table 1 shows a description of all model’s parameters and their preset values.

**Table 1.** Description of the parameters involved in model 1 (see [1, 26] for sources).

| Parameter | Description | Value | Units |
| --- | --- | --- | --- |
| $\beta(t)$ | Effective contact rate | estimated | days<sup>-1</sup> |
| $\psi$ | Effective vaccination rate | variable | days<sup>-1</sup> |
| $\phi$ | Vaccination immunity rate | 0.005556 | days<sup>-1</sup> |
| $\omega$ | Natural immunity rate | 0.005556 | days<sup>-1</sup> |
| $\gamma$ | Incubation rate | 0.196078 | days<sup>-1</sup> |
| $\delta$ | Asymptomatic recovery rate | 0.142857 | days<sup>-1</sup> |
| $\eta$ | Symptomatic recovery rate | 0.071429 | days<sup>-1</sup> |
| $\kappa$ | Reported recovery rate | 0.1 | days<sup>-1</sup> |
| $\nu$ | Screening rate | estimated | days<sup>-1</sup> |
| $\mu$ | Death rate | estimated | days<sup>-1</sup> |
| $\rho$ | Proportion of asymptomatic | 0.2 | % |
| $\epsilon$ | Proportion of symptomatic recovered | 0.85 | % |
| $\alpha$ | Proportion of reported recovered | estimated | % |
| $q$ | Asymptomatic infectiousness reduction | 0.45 | % |

### 2.2 Effective contact rate estimation

The main feature of the model is the time-dependent effective contact rate *β*(*t*). This function is defined by a piecewise cubic Hermite interpolating polynomial [13]. We set *b*_*k*_ as the effective contact rate at the *k*-th time change point, i.e. *β*(*t*_*k*_) = *b*_*k*_, *k* = 1, …, *K*. Fig. 3 shows a diagram of the structure of *β*(*t*). Hermite interpolation is used to guarantee that *β*(*t*) remains positive for all times. We emphasise that, instead of choosing the change times *t*_*k*_ equally spaced along the observation period, they are located at predetermined calendar dates associated to superspreding events, government interventions or other events that could affect the evolution of the epidemic curve.

**Fig. 3.**
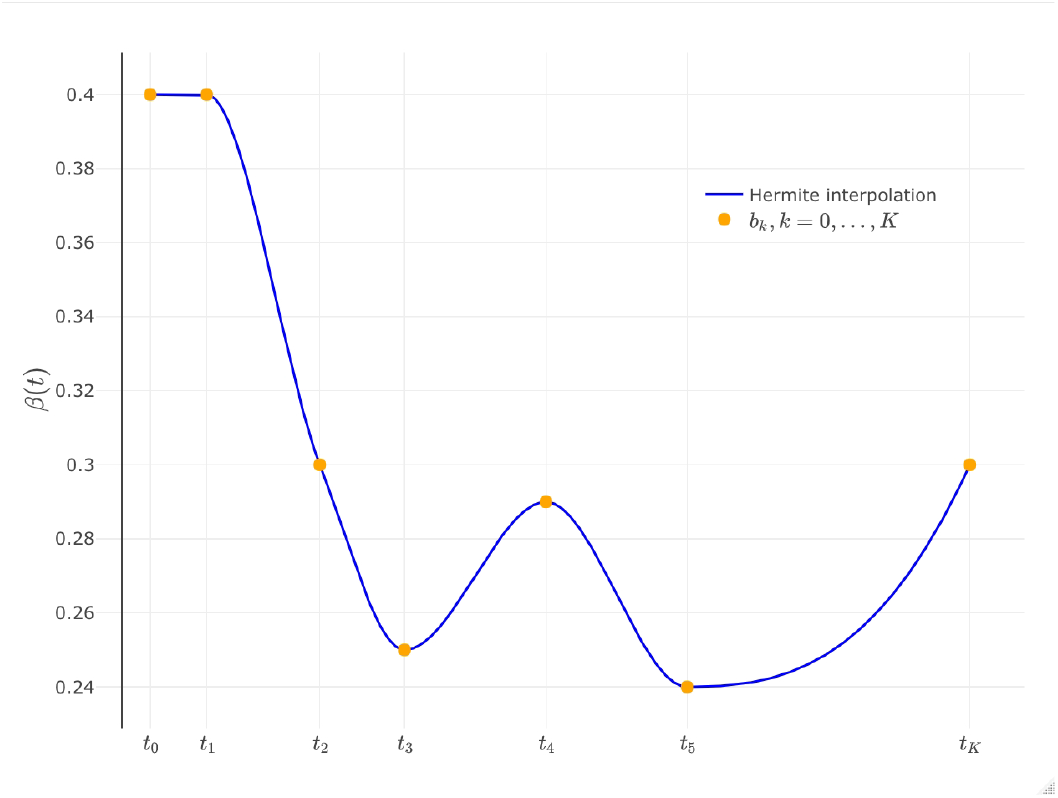
Construction of the effective contact rate *β*(*t*). It is defined by a piecewise cubic Hermite interpolating polynomial, parametrized by *K* change points at fixed times *t*_1_, *t*_2_, …, *t*_*K*_ and their corresponding values *b*_1_, …, *b*_*K*_.

The function *β*(*t*) (parameters *t*_1_, *t*_2_, …, *t*_*K*_, *b*_1_, *b*_2_, …, *b*_*K*_) must be estimated from data. We use federal national records from daily reported cases and deaths to create point and interval estimates of these parameters, within a Bayesian inference framework. Details about the inference can be found in Appendix A.

### 2.3 Projected scenarios

Our main point is that the effective contact rate *β*(*t*) in 2020 and early 2021 provides information that can be used to create scenarios for the evolution of the epidemic curve for the rest of 2021. There exists key calendar events or fixed dates where the Mexican population has family gatherings, political or civic activities every year, that during 2020 were associated to heightened transmission, heightened hospital demand, and increases in deaths. If this observation holds then, there are several ways in which we can project trends and define scenarios. One way is by simply prolonging for a few weeks or months the last observed trend of *β*(*t*) and see the effect on the number of cases and deaths using the model. This is a common approach to create short-term forecasts. It usually works as long as that last observed trend does not change in the data, which could occur in a few days, weeks, or even months. However, we can improve this methodology using our estimation of *β*(*t*): it is possible to know which dates are associated with changes in the trend and the magnitude of the contact rate. Therefore, we can assume that the percentage change observed in *β*(*t*) between two consecutive dates will be repeated in the same period the next year. To exemplify this idea, suppose that *β*(*t*) on December 24, 2020, was 0.2 and on December 31, 2020, was 0.3, implying that the contact rate increased by 50%. Then, to predict the change in the same period of 2021, the estimated contact rate on December 24, 2021, lets say 0.05, will increase a 50% to reach 0.075. Starting this process on the last date of the estimated contact rate, it is possible to generate projections that incorporate the history of the epidemic.

## 3 Results

To exemplify this methodology, the epidemic data of several states of Mexico will be used to estimate the effective contact rate *β*(*t*) and then to propose scenarios for the rest of 2021 and early 2022.

### 3.1 Key superspreading events

The overall perspective of the epidemic management in Mexico can be fairly summarized by saying that a) all non-pharmaceutical interventions have been and are, to date, non-mandatory but voluntary except for some policies regarding closure or reopening of businesses; and b) the population increased mobility and social activities during certain periods is strongly associated to nation-wide holidays. The list of dates that we consider important for every state is given in Table 2.

**Table 2.** Key calendar dates associated to superspreading events used as change points for the effective contact rate *β*(*t*) in the state of Tamaulipas. Except for the date of the first COVID-19 case (by symptoms onset), all the other dates are the same for each state analyzed. Mexico City reported its first case on February 20, 2020 and Querétaro state reported its first case on March 5, 2020.

| Date | Description | Date | Description |
| --- | --- | --- | --- |
| 2020-03-13 | First COVID-19 case | 2020-12-24 | Christmas |
| 2020-03-23 | NPI start | 2020-12-31 | New year's eve |
| 2020-04-30 | Children's day | 2021-01-06 | Wise men day |
| 2020-05-10 | Mother's day | 2021-02-14 | Valentine's day |
| 2020-06-21 | Father's day | 2021-03-14 | Benito Juarez birth |
| 2020-09-16 | Independence day | 2021-04-04 | Easter |
| 2020-11-02 | Day of the Dead | 2021-04-30 | Children's day |
| 2020-11-21 | Buen Fin (Black Friday equivalent) | 2021-05-10 | Mother's day |
| 2020-12-12 | Day of the Virgin of Guadalupe |  |  |

To provide evidence on the key calendar dates superspreading hypothesis, several indicators are analyzed such as positivity, hospitalizations, mortality, among others. Only positivity is presented but other indicatores can be found in the supplementary material. When analyzing raw counts of the number of cases by day, the variability between days makes it difficult to determine the behavior of the data curve. To deal with this problem, we use a simple moving average with a window size of seven days, making it easier to identify overall trends. The positivity rate is defined as the ratio of positive tests to suspected cases.

Figure 4 shows the positivity rate for three states: Mexico City, Queretaro and Tamaulipas. Estimations are noisy but still it can be appreciated that, depending on the length of the holiday, there is an increase either immediately after or even during the key event. Also, changes in the trend can be associated to several key dates. As an example, December celebrations such as Christmas cause an increase in the positivity rate, which starts decreasing after New Year.

**Fig. 4.**
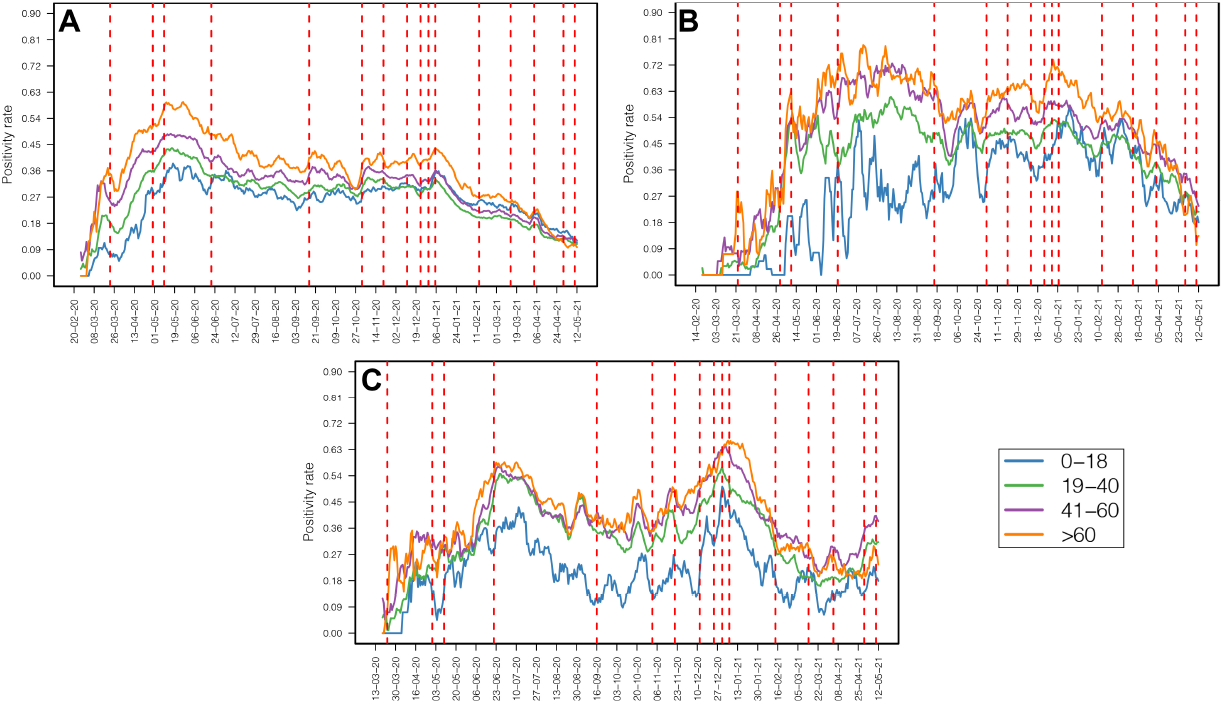
Positivity rate by age group for A) Mexico City, B) Queretaro state and C) Tamaulipas state. Dashed red vertical lines give the dates listed in Table 2.

It must be pointed out that changes in these indicators are not exclusive to the key dates we have selected (Table 2). Also, not necessarily all the dates chosen have the same impact for each state. However, our main hypothesis is that significant changes are indeed associated to the dates previously selected and they are enough to drive the general epidemic trends in each state.

### 3.2 Effective contact rate estimates for Mexico

As mentioned in Section 2.2, the effective contact rate is determined by an interpolating function and parametrized by (*t*_0_, *t*_1_, …, *t*_*K*_, *b*_0_, *b*_1_, …, *b*_*K*_). Times *t*_0_, …, *t*_*K*_ are determined by the key dates in table 2. We point out that *β*(*t*_0_) and *β*(*t*_1_), *b*_0_ and *b*_1_ respectively, are equal. Given that *t*_1_ represents the time then the first mitigation measures were implemented, setting *b*_0_ = *b*_1_ implies that the effective contact rate is constant right before the mitigation interventions that altered the natural progression of the epidemic. All other parameters *b*_1_, …, *b*_*K*_ are estimated. It is assumed that, at the beginning of the pandemic, there are no vaccinated individuals, no reported infected individuals, no recovered cases and no deaths. The number of exposed, asymptomatically infected and symptomatically infected individuals are also estimated. Finally, the COVID-19 death rate, the screening rate, and the proportion of reported recovered individuals can vary from one state to another and therefore are also estimated. See Appendix A for more details.

Fig. 5 shows the point-wise posterior median estimate of *β*(*t*) for several states in Mexico, including the three examples previously discussed: Mexico City, Queretaro and Tamaulipas. Although each place has a different curve there are, nonetheless, common patterns. Notice that the rate remains low between June 2020 and September 2020 and the effect of the winter holidays is clear in each state. It is interesting that a few states showed an increase in the contact rate after the start of mitigation measures such as Queretaro, Guanajuato and Campeche. Overall, we can group states that show very similar behavior, a type of information that could potentially be used to classify epidemic curves. This will be explored in a forthcoming work.

**Fig. 5.**
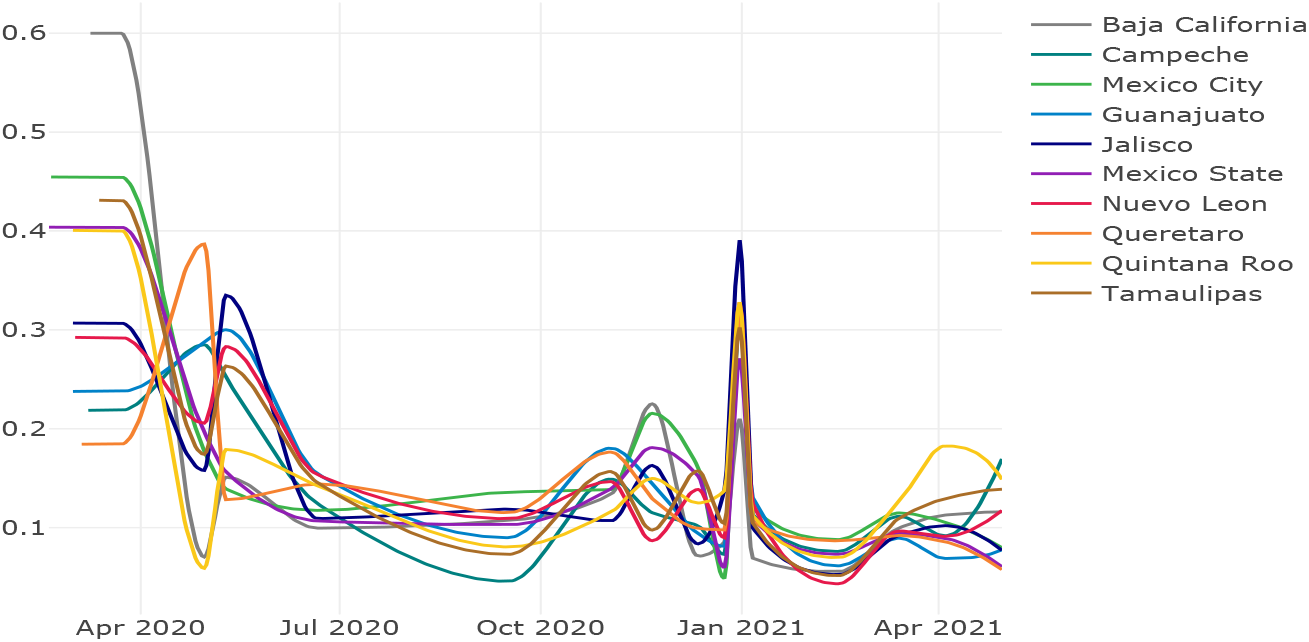
Effective contact rate *β*(*t*) estimated for several Mexican states from the beginning of the pandemic (late February to early March) in 2020 until April 30, 2021.

### 3.3 Scenarios

As described above, we modify the last estimate of the effective contact rate, *β*(*t*), corresponding to April 30, 2021, with the purpose of generating projections supported on the effective contact rate patterns observed on the same dates in 2020. However, the one aspect of the epidemic that is clearly different from last year’s is the availability of vaccines, and thus this factor requires especial handling. Vaccination in places like Mexico is difficult to model since many different vaccines are applied, each with very different characteristics, such as the ones produced by Pfizer, Moderna, AstraZeneca and CanSino, to mention some. Moreover, the national vaccination strategy contemplates several stages depending on the stock of vaccines, which at the moment is very limited. By the end of May 2021, 16% of the population has been vaccinated, although only 9% has a complete vaccination scheme [20]. Nationally, vaccination started at the end of December 2020, but there are differences in the vaccination roll out in each state and limited information about it. For the purposes of this work, it is assumed that vaccination started on February 15, 2021, which is the date when most of the states started vaccination for the general population above 60 years old. The efficacy of the vaccines is set to 0.95 and that the expected coverage of the vaccines is 30%, 50% and 70% of the population after the first year. These three scenarios are considered to analyze the effect of vaccination.

Importantly, the variability of the estimates of *β*(*t*) is high, especially at the beginning of the epidemic. This is mainly caused by the initial conditions that are also estimated. Kermack-McKendrick models are very sensitive to changes in these conditions. As a consequence, a subset of projections for some states is extremely high and biologically unfeasible as they imply very high values of the effective reproduction number *R*_*t*_. For that reason, the predictions shown in this work are limited to those with estimates of *R*_*t*_ for the prediction period below the maximum observed *R*_*t*_ in the past. We think this restriction is reasonable.

Fig. 6 shows the results for Tamaulipas state. The model correctly describes the main trends during the estimation period. The effect of the vaccination is to decrease the number of new reported cases and deaths, although qualitatively, the behavior of the predictions is the same. Thus, if similar conditions to 2020 occur in 2021, the most likely scenarios indicate that the number of new infections will be low and decreasing for the rest of the year. Nonetheless, there is still the possibility of an important increase of the number of infections in the following months.

**Fig. 6.**
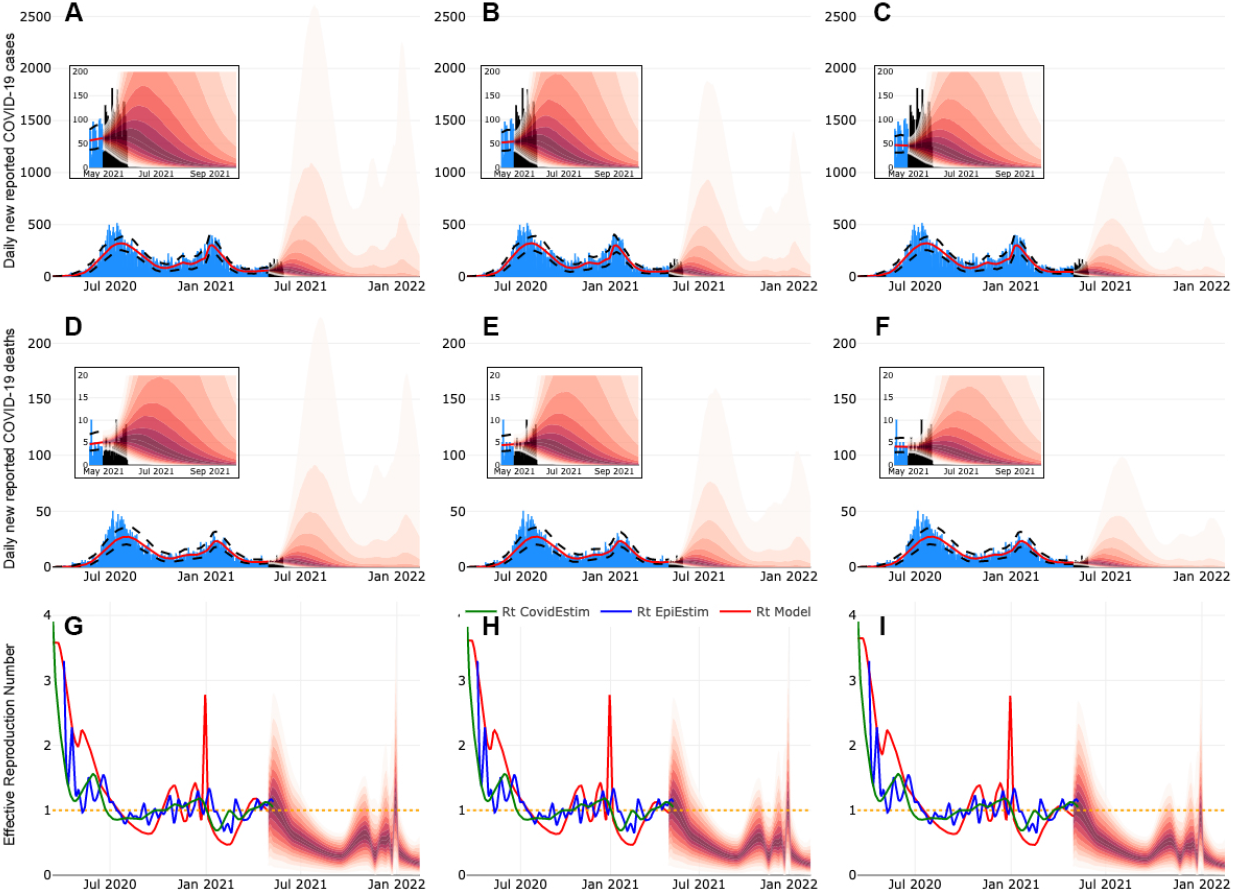
Projected scenarios for Tamaulipas state. First, second and third columns show scenarios when target vaccination coverage are 30%, 50% and 70%, respectively. These coverages are achieved in a year. The insets show an enlarged view of the first months of each projection.

Fig. 7 shows the scenarios for Mexico City. In this case the most likely trend is that the number of cases and deaths decrease in time with very low levels on infections for the rest of the year. There is a small probability of an outbreak at the end of 2021 and the beginning of 2022, and its magnitude could by similar to the one observed for the same period in 2020 if only 30% is vaccinated by that date. Fig. 8 shows the scenarios for Queretaro state. In this case the number of cases and deaths go down very fast and there will be no more outbreaks.

**Fig. 7.**
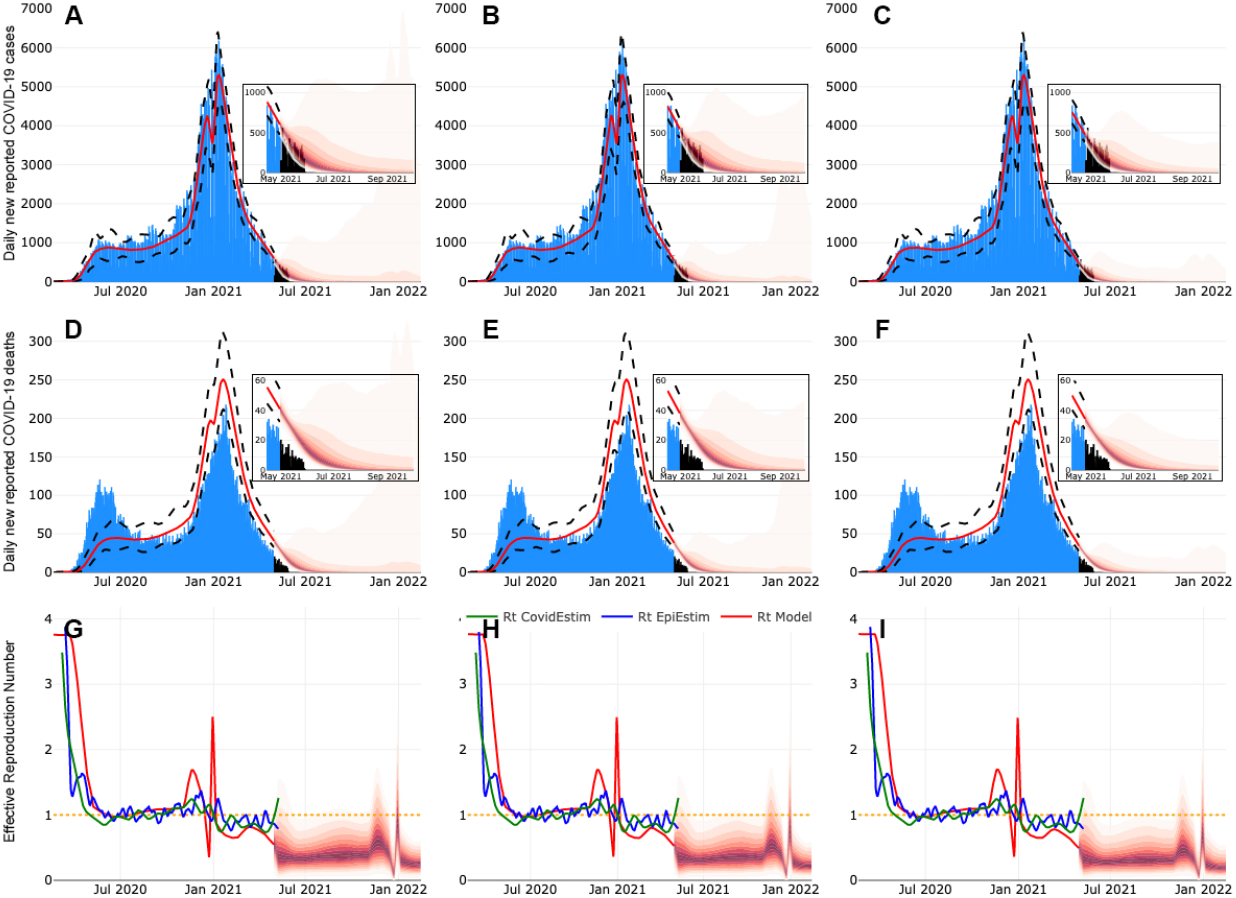
Projected scenarios for Mexico City. First, second and third columns show scenarios when target vaccination coverage are 30%, 50% and 70%, respectively. These coverages are achieved in a year. The insets show an enlarged view of the first months of each projection.

**Fig. 8.**
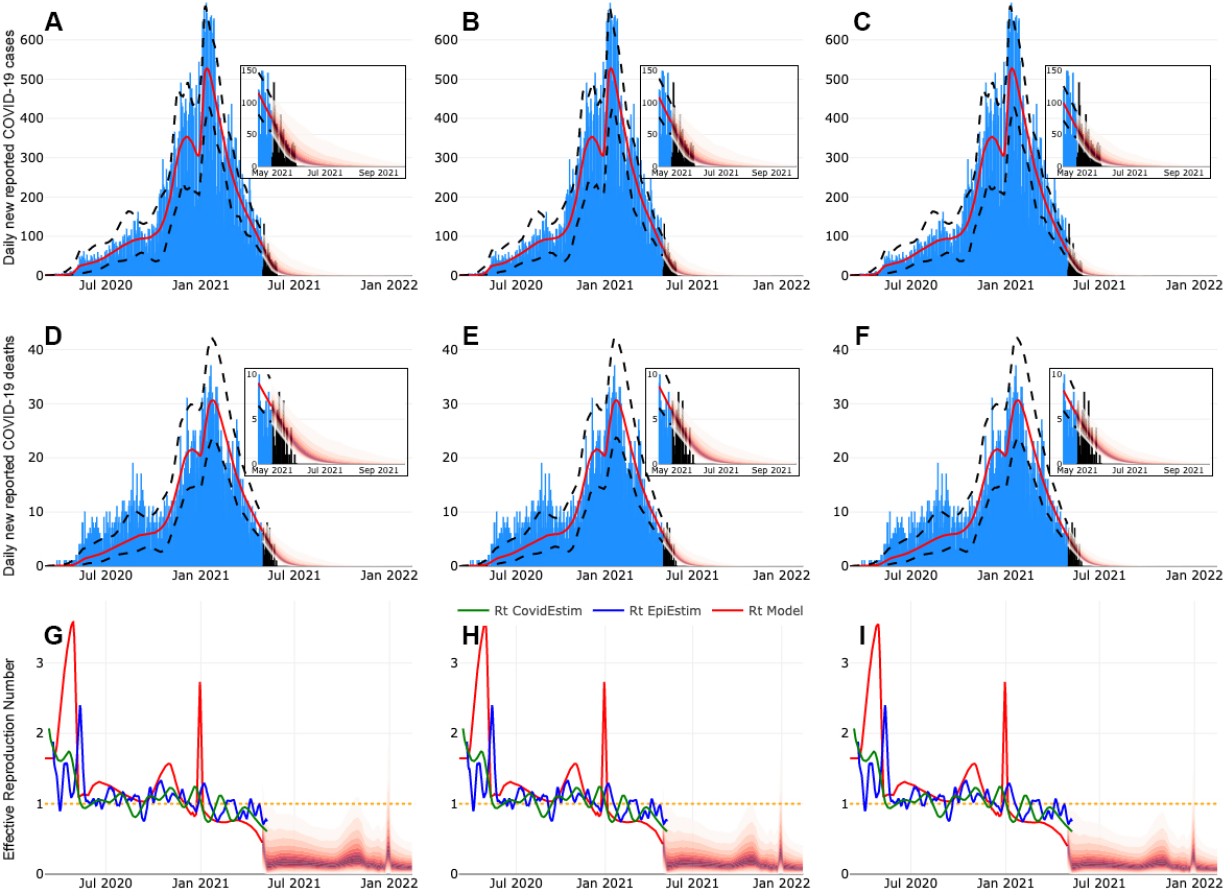
Projected scenarios for Queretaro state. First, second and third columns show scenarios where target vaccination coverage are 30%, 50% and 70% during the first year, respectively. The insets show an enlarged view of the first months of each projection.

Black bars in Figs. 6, 7 and 8 show two weeks of available data on reported cases and deaths for the last three weeks that was not used for in the estimation. Projections agree with the observed data. These figures also show 3 estimates of the effective reproduction number for each scenario according to three different methodologies, one derived form the equation 3 and the other two obtained using EpiEstim [6, 30] and Covidestim [10, 22].

## 4 Discussion

Our objective is to generate a range of scenarios using the known history of the epidemic, that could be useful to evaluate its possible evolution and its likely impact on incidence and mortality. The fundamental assumptions behind this methodology are that the effective contact rate incorporates the main superspreading transmission events during last year, that each region has an independent epidemic not explicitly interconnected with other regions and, finally, that there are no new highly transmissible variants active during the timeline of the forecasts.

The magnitude and changes of the contact rates on key dates on 2020 together with the magnitude of *R*_*t*_, constitute two indicators that provide an approximation of what could happen during 2021. The main assumptions are that these dates will again constitute superspreading events and that the evolution of the epidemic curve in 2021 will still be highly influenced by the same events. The idea is simple but the use of key calendar dates creates a practical basis, in the absence of significant testing and contact tracing, to anticipate changes in the transmission of the disease that can provide mid to long term outbreak risk projections. There is no doubt that superspreading events will still be important for the COVID-19 pandemic however, it is not clear what the magnitude of their impact will be. The methodology presented here is a step towards the solution of this problem.

It could be argued that this procedure will not be useful because the conditions of the previous year will not be the same for the next one. However, in places like Mexico the proposed procedure is sensible because: 1) lockdowns and other non-pharmaceutical interventions are not mandatory, 2) there is very limited testing and practically no contact tracing, and 3) significant superspreading events occurred mostly during holidays that happen every year. Nonetheless, we are aware that, for now, only the short-term predictions can be evaluated.

New events that were not present during 2020 can have an important impact in the evolution of the epidemic curve during 2021. For example, on June 6, 2021, there will be Federal Elections in the country, which could potentially constitute a superspreading event. If that were the case, the current projections may fail. However, it is possible to adjust the estimates to accommodate this date. The process will be less immediate than the one used here because there were no elections during 2020. Nonetheless, scenarios can be created based on the changes observed in other superspreading events. As was mentioned before, there are several ways of using the information contained in *β*(*t*).

Our model does not explicitly introduces seasonality that could start acting on the epidemic by October 2021 under the assumption that COVID-19 behaves similarly to influenza. However, since the pattern of changes of the observed contact rate for 2020 is being used, any seasonal effect with impact on last year’s contact rate will be inherited into the projected contact rates of 2021.

Even without the projections, the estimation of a time dependent effective contact rate alone provides interesting information. The evolution of the epidemic curves of different locations can be compared, and possibly grouped, using these estimations. In the case of Mexico, a common feature to all states is that the effective contact rate had one fast period of growth on the winter holidays (December 12 to December 31, 2020), supporting the idea that superspreading events did occur in the proposed dates. Even when not all the key dates considered in the analysis have an significant effect in all the states, the results suggest that it is important to closely look at the behavior of the epidemic on these dates.

Finally, this study highlights the importance of enforcing the application of NPIs in Mexico and countries of similar characteristics. In all our scenarios, the reduction in *β*(*t*) can be achieved only through them, given the low vaccination rate that exists at the end of May. This is not surprising, a recent study [23] has shown that even with high efficacy vaccines and a greater coverage than the current one in Mexico, vaccination alone cannot achieve a significant reduction in cases if NPIs are relaxed too quickly.

## Data Availability

All the data used in this work is publicly available.

https://www.gob.mx/salud/documentos/datos-abiertos-152127

## Acknowledgements

JXVH and MSC acknowledge support from UNAM PAPIIT DGAPA IN115720 and IV100220 grants. CERHV and RHM are grateful for the support of UNAM PAPIIT grant IG100221. MAAZ acknowledges support from PRODEP Programme (No. 511-6/2019-8291).

## Author contributions statement

MSC, MAAZ and JXVH conceived the idea; CERHV and RHM analyzed data, MSC and MAAZ performed the model projections and parameter estimation, all authors discussed, reviewed and wrote the paper.

## Additional information

Further analysis for other Mexican states is shown in the site: https://github.com/ProjectsEpi/KeySuperspreadingEvents.git

## A Statistical inference

In order to create predictions for the evolution of the COVID-19 pandemic during 2021, we focus on the estimation of the time dependent contact rate *β*(*t*) during 2020 and the start of 2021 as was described in Section 3. To simplify the estimation process, *β*(*t*) is assumed to be a piecewise Hermite interpolating polinomial that only changes at preset times *t*_1_, *t*_2_, …, *t*_*k*_, and it is constant before *t*_1_. Then, instead of estimating a continuous function, we only need to estimate the values of the effective contact rate *b*_1_, *b*_2_, …, *b*_*k*_ at *t*_1_, *t*_2_, …, *t*_*k*_.

The dates where the effective contact rate changes are described in Table 2. Those dates are converted into a numeric scale to get *t*_1_, *t*_2_, …, *t*_*k*_ simply by calculating the number of days from the first reported case (by symptoms onset). For each Mexican state, the starting date for the analysis is different.

The initial number of exposed (*E*_0_), asymptomatic (*I*_0_) and symptomatic (*Y*_0_) individuals will also be estimated as these are important unknown quantities. Parameters *α, µ* and *ν* depend on the state analyzed. Parameter 1*/µ* is directly estimated from the records of confirmed cases by calculating the average difference in days between the date of symptoms onset and the date of death, while 1/*ν* is calculated as the average difference in days between the date of symptoms onset and the date of hospital registration. Only parameter *α* is included in the inference process.

Let ***θ*** = (*E*_0_, *I*_0_, *Y*_0_, *α, b*_1_, *b*_2_, …, *b*_*k*_) the vector of parameters that will be estimated using a Bayesian inference approach. All the other parameters needed to solve model (1) are fixed and their values can be found in Table 1.

Let *X*_*j*_ and *Y*_*j*_ be the random variables that count the number of daily COVID-19 reported cases and deaths at time *t*_*j*_, respectively, for *j* = 1, 2, …*n*, where *t*_*j*_ represent the number of days since the first reported case by symptoms onset. We assume that the probability distribution of *X*_*j*_ and *Y*_*j*_, conditional on the vector of parameters ***θ***, is a Poisson distribution such that

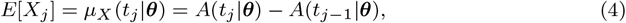

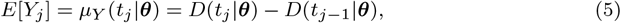

with *A*(*t* |***θ***) ad *D*(*t* |***θ***) being the cumulative number of reported cases and deaths according to model (1). Assuming that variables *X*_1_, *X*_2_, …, *X*_*n*_, *Y*_1_, …, *Y*_*n*_ are conditionally independent, then the likelihood function is given by

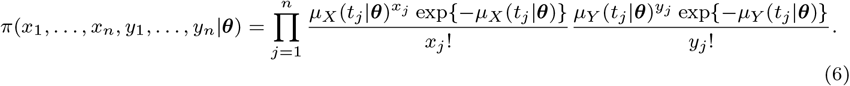

The joint prior distribution for vector ***θ*** is a product of independent probability distributions. For all the initial conditions *E*_0_, *I*_0_, *Y*_0_, the prior distribution is Uniform(0,20) and for all the contact rates *b*_1_, …, *b*_*k*_ the prior is Uniform(0,5). The prior for *α* is a Beta distribution with parameters *c*_1_ = 20.0 and *c*_2_ = 2.2 which has an expected value of 0.9. Then

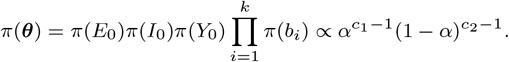

The posterior distribution of the parameters of interest is

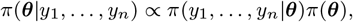

and it does not have an analytical form since the likelihood function depends on the numerical solution of the ODE system (1). We analyze the posterior distribution using an MCMC algorithm called *t-walk* [11]. For each state, three chains of 1,000,000 iterations are run, from which 100,000 are discarded as burn in. At the end, only 2000 iterations are used to create the estimations presented in this work. The posterior predictive distribution is used to create the scenarios for the rest of 2021 under the three specific values of the vaccination rate *φ*.

